# Mental health services implementation in Colombia – a systematic review

**DOI:** 10.1101/2023.01.17.23284625

**Authors:** Germán Andrés Alarcón Garavito, Rochelle Burgess, María Cecilia Dedios Sanguinetti, Laura E.R. Peters, Norha Vera San Juan

## Abstract

Colombia’s mental health services have a complex history shaped by 60 years of armed conflict, a predominantly clinical approach to mental health, and social factors such as inequities and stigma. The 1990 Caracas declaration proposed a shift towards decentralised community mental health services and interventions based on the recovery approach and emphasis on social determinants of mental health in the Americas. Colombia has adopted these approaches in its legal and practical framework in recent years, but implementation has been uneven. This systematic review aims to contribute to mental health services understanding in Colombia by reviewing published studies about mental health services and interventions.

A search was conducted to explore peer-reviewed studies on Colombian mental health services across five databases (Medline, PubMed, Scopus, Scielo and BVS [Biblioteca Virtual de Salud in Spanish]) on papers published in the last ten years and without language restrictions. The Consolidated Framework for Implementation Research (CFIR) was used to structure the analysis and identify barriers and facilitators during the implementation of mental health services. We adapted the CFIR to attend to gender, race and age informed by the Socio-Political Economy of Global Mental Health framework, given the importance of these factors to the Colombian health landscape.

1 530 records were identified, and 12 articles met all inclusion criteria and were included in the analysis. 8 papers described substance use disorders services, 11 involved multidisciplinary healthcare professionals, and 7 were implemented at a local scale. The primary barriers to implementation were the lack of coordination, high workloads, and low funding. Facilitators included the use of protocols, and the involvement of communities, stakeholders, users, and external champions.

Findings suggest the continued importance of community and recovery approaches and efforts to improve coordination between multi-sector actors involved in the mental health spaces (e.g., public, and private organisations, users and their families).

## Introduction

In the last 30 years, mental health systems and services have received more attention and urgency to prioritise by governments. Global crisis such as the COVID-19 pandemic has soared pressure on people’s mental health and has risen general mental health awareness. Nevertheless, how governments and mental health systems should respond is still a general debate. For instance, Colombia has advanced in building a relative robust health policy space to address these challenges. However, translating visions and concepts into real-world changes tends to be a convoluted pathway full of barriers and enablers, which should be avoided and supported respectively to achieve the goals and objectives articulated in policy documents (1).

### Global and regional situation

In 2007, The Lancet launched the first global mental health series outlining the treatment gaps in low-and-middle-income countries (LMICs) and the necessity of better cost-effective mental health interventions (2). Furthermore, the ratification of the Sustainable Development Goals in 2015 included the prevention and promotion of mental health with a strong emphasis on substance abuse, including the use of alcohol (3).

The WHO Mental Health Action Plan 2013-2020 led many states along pathways to strengthen leadership and governance for mental health, increase the provision of mental and social care services and support better information systems for evidence and research (4). In Latin America, such efforts are predated by the *Caracas Declaration of Mental Health, and Human Rights* from 1990, which highlighted the necessity of adapting primary care lenses and community-based services in mental health. It also promoted more team planning and the reduction of the role of institutionalization in mental health services, expanding voices in mental health spaces (5).

Critiques against the global mental health agenda remain. Many report of the limited consultation with social scientists, people with psychosocial disabilities, victims of negative mental health services, and others (6). Moreover, analysts also argue that the proposed mental health models are limited by “Western” neo-colonial lenses, disregarding traditional and cultural understandings of mental health interpretations (7). This situation fosters the replication of services solely built on the biomedical model.

One of the consequences is that non-pharmaceutical interventions such as psychotherapy or community-based programs are actively overlooked. Also called “psychiatrisation,” the biomedical perspective favours pharmaceutical companies, governmental supporters behind them, and even some psychiatry and mental health practitioners. This scenario has facilitated a profitable world market for these companies and their stakeholders (8). The prioritisation of pharmacological treatment has also produced a normalisation of adverse effects of psychiatric medications, which are incredibly relevant and should be reported to regulatory agencies and manufacturers for quality and research purposes (9,10).

Furthermore, in Latin America, the outlook on mental health financing is complex. It is not a surprise that high-income countries (HICs) spend more on mental health than they do on other health problems. While HICs’ funding is around 5% of the total health budget, LMICs’ is between 2 and 9 times less. Overall, the percentage of the budget used on mental health services is not proportional to the negative impact that none or limited mental health provision generates in society, as stated by multiple experts in the last 20 years (11,12).

In the Americas in 2018, the treatment gap for mental disorders was up to 71.2% for any mental disorder and 65.7% for severe to moderate disorders. The difference between North and South America is evident, considering that the treatment gap for substance abuse is 83.7% and 69.1%, respectively. Some individual factors behind this reality are the lack of belief in treatment, financial barriers, and stigma, among others (13).

### Mental health policies and services in Colombia

Colombia has suffered decades of violence which has had a major impact on Colombians’ mental health (14). More than 60 years of internal conflict (1960) have fractured social structures and intensified the transmission of communicable and non-communicable diseases (14). War tactics like massacres, anti-personnel landmines, and gender-based and sexual violence drove internal displacement and the escalation of substance abuse (15). These circumstances can also trigger a plethora of mental health difficulties among victims (including legal and illegal armed forces, civilians, and foreigners) (14).

According to the National Health Institute of Colombia, the consequences of armed conflict on mental health include high rates of post-traumatic stress disorder (PTSD), depression, anxiety, and suicidal conduct (14). Additionally, the conflict directly affected human healthcare resources due to direct attacks on the workforce, causing a cessation in the training of new healthcare professionals (HCPs) in some regions such as Arauca, Guaviare, Caquetá and Putumayo, which were the regions with the highest indicators of violent conflict intensity (14). Healthcare services became increasingly urban-based due to the intensified violence that led to the reduction of healthcare centres in conflict-affected rural areas, and many people stopped accessing care due to mobility limitations and financial hardship (14).

In 1993, the National Law 100 determined that healthcare services must be administered and partially operated by healthcare insurers (Empresas Promotoras de Salud [EPS] in Spanish), and the population was divided into two income-based schemes; subsidised and contributory (16). Insurers hire providers (private practitioners, hospitals, clinics, laboratories) to deliver health services stated in the national health benefits packages (Plan de Beneficios en Salud (PBS) in Spanish) (17).

This law established a legal right to healthcare based on opportunity, quality, and comprehensive coverage. Healthcare coverage increased from 23% to 97% (18), with a benefits package affordable for most of the population (19). However, the law weakened mental health services, revoking programs like Day Hospitals, restricting access to psychotherapy after one month, and only covering early-stage treatment (19). Consequently, the number of legal actions (also known as *“Tutelas”* in Colombia) against healthcare insurers skyrocketed. These oversights required a range of reforms, new policies, and amends.

One of these reforms was the first version of the National Mental Health Policy in 1998, strengthened with the Mental Health Law in 2013 (20,21). These legal and strategic frameworks aimed to improve access, quality, and coverage. However, their local implementation has been limited and insufficient (12). According to Holguín and Sanmartín-Rueda (2018) (23) current mental health services in Colombia are centred on financial profitability for insurers, and due to weak inspection and regulation, numerous deficiencies in the flow of financial assets, and corruption lawsuits, the legal right to healthcare has been undermined.

In parallel at the social level, stigma, which compromises service quality worldwide, has been categorised as one of the most notable factors that prevent accessing mental health services in Colombia (24). According to Campo-Arias, Oviedo and Herazo (2014) (26) the most common sources of stigma are the surrounding community, family, employees and colleagues, and media. People believe that the only causes of mental disorders are substance abuse, brain disease, and spiritual reasons such as evil spirits or “God’s punishments”. This lack of additional understanding may be due to a deficiency of available information regarding mental health disorders’ nature and multiple causes.

On the whole, there is a global and regional space with ideas on mental health approaches, and Colombia is aiming to develop laws and policies influenced by these ideas and supported by massive surveys and monitoring actions such as the National Mental Health Survey in 2015 or the 2021 mental health statistical reports by the National Statistical System (Departamento Administrativo Nacional de Estadística or DANE in Spanish). Thus, both the National Mental Health Law and Policy have represented an opportunity for Colombia to develop integrated mental health models and frameworks based on national evidence, and community approaches and consider the social determinants of mental health (SDMH). However, multiple studies, independent reports and civil complaints spell out the decay of mental health services and how they are failing to satisfy mental health care needs (27–33), demonstrating that this supportive policy environment does not automatically translate into effective and accessible services in practice.

### Aim

Despite the robust mental health policy environment in Colombia, there is a gap on why implementation failures remain. The purpose of this paper is to identify the barriers contributing to these gaps in delivering effective and accessible services aligning with policy ambitions. It also aims to contribute to existing knowledge by identifying the facilitators for implementing mental health services in Colombia.

The following research questions guided this review:

1. What are the reported mental health services in Colombia?

2. What are the main challenges with the implementation of mental health services?

3. What are the facilitators of the implementation of mental health services?

## Methods

### Design and approach

This systematic review included chronological identification, screening, selection and synthesis (34,35), and followed a systematic approach to ensuring that all the available information was incorporated (35). This systematic review was conducted following the Preferred Reporting Items for Systematic reviews and Meta-Analyses (PRISMA) guidelines. No formal protocol was developed or published for this review.

### Implementation science framework

The relevance of evidence for decision-making in policy is well known, especially in healthcare. Healthcare settings are usually complex environments, with limited resources and multiple actors, behaviours, and structures that complicate taking decisions (36). While the production of evidence rises every day, it is a well-known fact that without methodical adoption processes for both clinical and non-clinical interventions, significant changes in health outcomes cannot be expected (37). Therefore, it is crucial to determine whether evidence can be implemented and what impedes its acceptance.

Implementation science’s scope goes beyond individuality, considering the context and structure of the settings where the implementation is operating (38). Some vital elements of implementation sciences are the frameworks and theories involved and their evaluation processes (36).

To increase the comparability of the results of this review, we used the CFIR. It comprises 39 different constructs divided into five domains: intervention characteristics, outer setting, inner setting, individual characteristics, and process (39). The CFIR was chosen because it provides a high-quality and integrative tool to assess implementation processes in multiple disciplines. In English et al., (2011) (41) this tool proved to be valuable to guide implementation assessments in an under-funded setting in Kenya, comparable to the Colombian settings. Nevertheless, the same study acknowledged that not all the constructs can be used in all scenarios due to a lack of available data or the reduced applicability of domains or constructs. Likewise, for this work, we only used the appropriate constructs from four domains, which are defined in table 1.

**Table 1.**
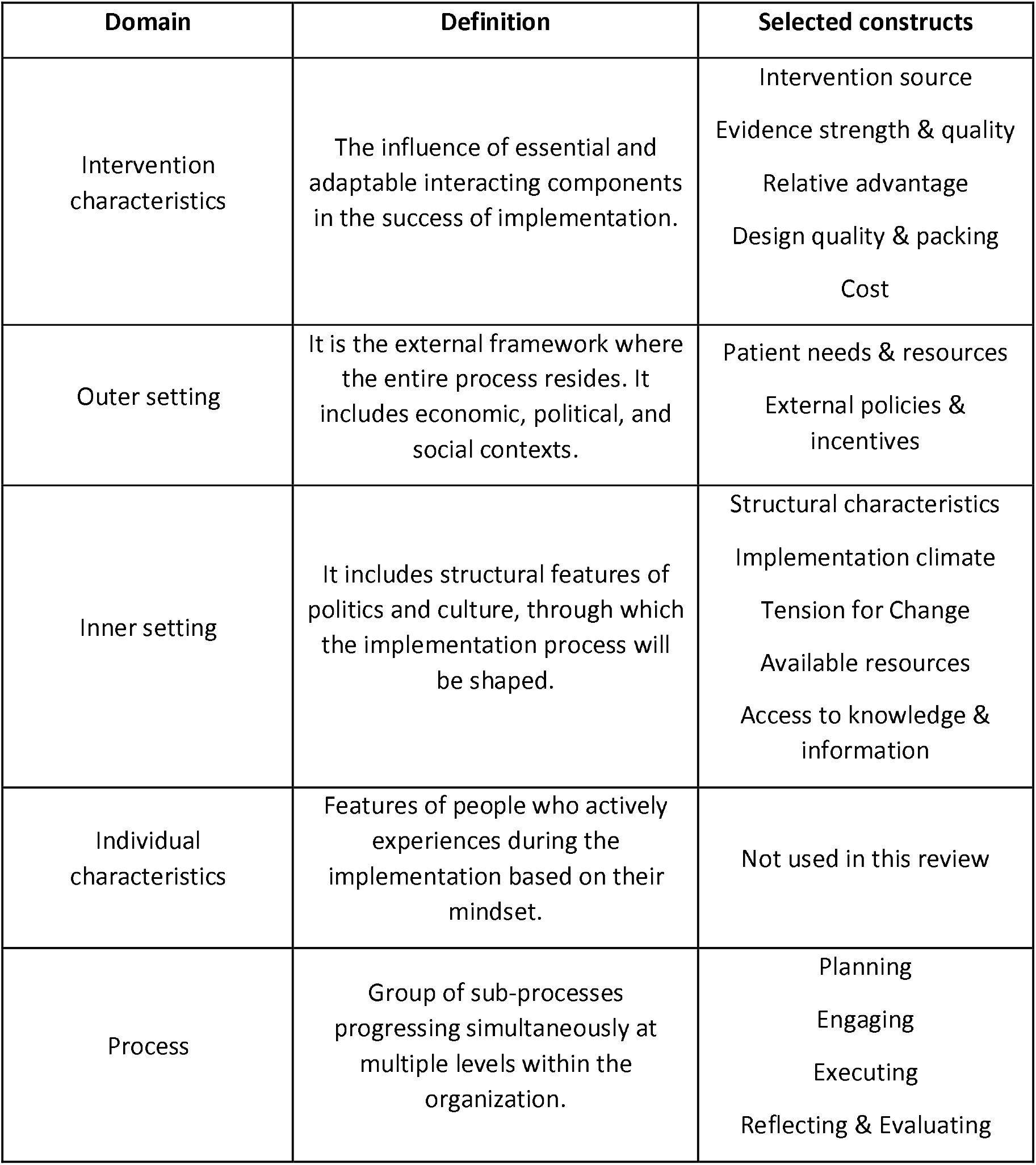
Overview of the CFIR.

The domain of characteristics of individuals was not used since very little data from included studies was about individuals’ beliefs, features, or capabilities. Furthermore, as the CFIR is focused on institutions and organisational problematics during implementation, factors such as religious beliefs, gender, ethnicity, or race are not actively considered within the framework despite their significant influence on mental health services. To cover these missing factors, we adapted the CFIR adding a new domain of socio-political economy in mental health based on the Socio-Political Economy of Global mental health framework proposed by Burgess et al., (2022) (43)

Specifically, we included three of their nine factors: I. Gender and sexuality, II. Racism, caste, xenophobia, and III. Age and disability. These factors were selected because they allowed us to explore their influence on determining mental health, their representation in the literature, and the existence of relevant information. We did not include other proposed factors such as religion, spirituality, or neighbourhood dynamics due to a foreseeable lack of information, while political dynamics were not incorporated since this kind of information was covered by the CFIR (domain of outer setting, construct of external policies and incentives).

### Search strategy

#### Databases

The literature search was performed between June and July 2021 using five databases: Medline (OVID), PubMed, Scopus, Scielo and BVS (Biblioteca Virtual de Salud in Spanish). These databases were selected because of the amount of peer-reviewed available information, and in the case of Scielo and BVS, due to their inclusion of Latin American literature.

#### Search equation

The search terms were established based on MeSH (Medical Subject Headings) terms and the Spanish counterpart DeCS (Descriptores en Ciencias de la Salud in Spanish) for mental health. Search terms included relevant words to refer to mental health and Colombia. It was necessary to use broad search terms because key literature results did not appear during preliminary searches. Detailed searching terms and combination strategies are described in **S1 Appendix 1. Search strategy**.

Location terms included the names of the five principal cities with more than one million inhabitants in Colombia. These cities were incorporated in the searches done in Scopus, Medline (OVID) and PubMed because it was noticed during earlier search checks that multiple possible eligible studies did not mention the country but specifically a city. It was also noted that using this strategy with the Boolean term “OR” did not risk losing data because studies mentioned at least the country or a city. This strategy was unnecessary in either Scielo or BVS since adding city names significantly lost results.

The search was limited between 2011 and 2021 to capture the most current state of mental health systems in Colombia while still capturing significant modifications of mental health policies (e.g., The Decennial Public Health Plan in 2012, National Mental Health Law in 2013), and the first years after the sign of the peace agreement in 2016 (44). Articles were screened at the title and abstract level using *Rayyan* software (45) based on the specific inclusion and e**xclusion criteria presented in Table 2**.

**Table 2.**
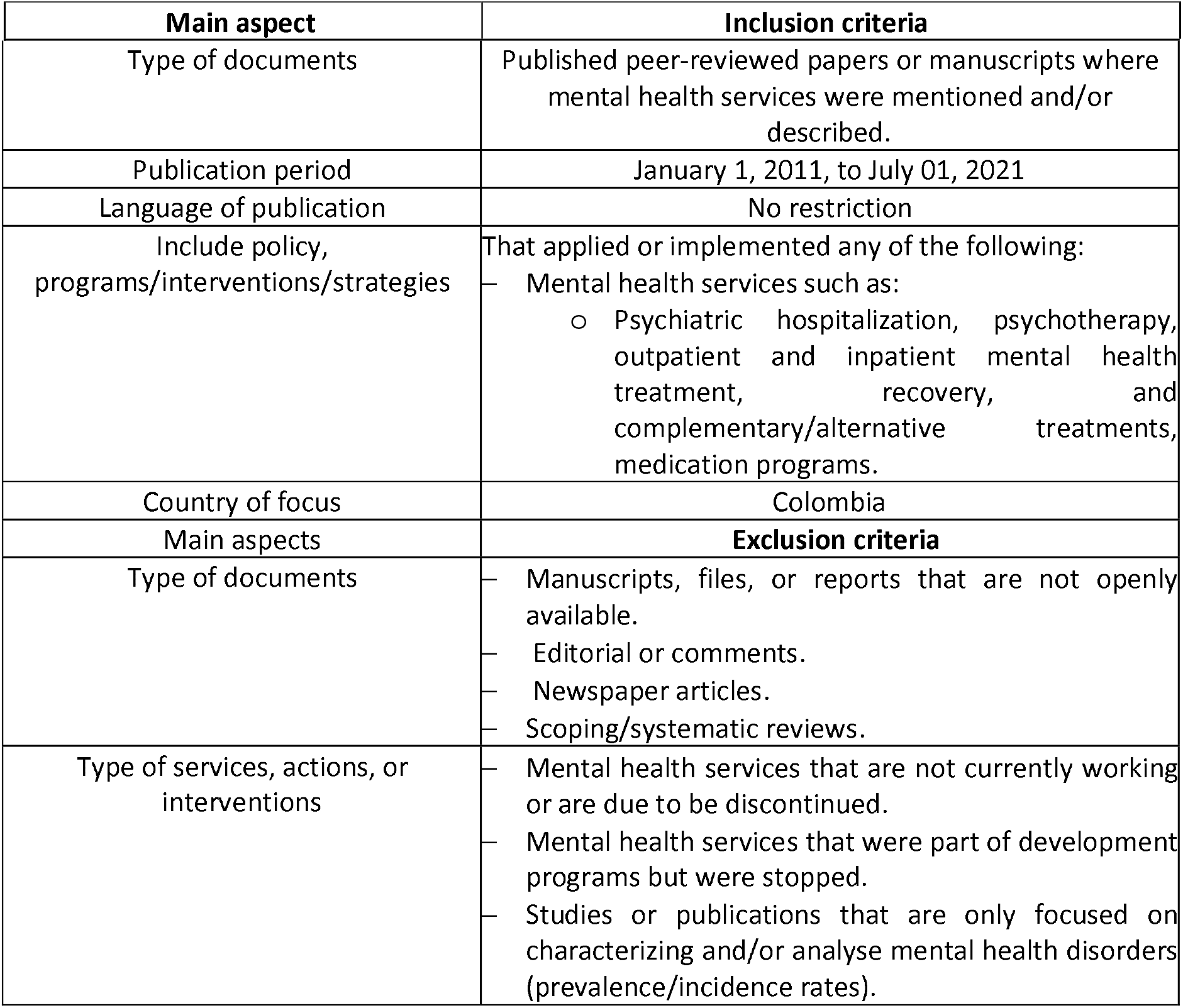
Inclusion and exclusion criteria.

## Data extraction

All the information was charted in an *Excel* data extraction form developed following the research objectives and based on the CFIR and the Socio-Political Economy of Global Mental Health framework.

Data included basic information (authors, year) and identification (e.g., mental health problem addressed, level of healthcare service, human resources, and population, among others). This review classified interventions into psychiatric hospitalisation, psychotherapy, outpatient and inpatient mental health treatment, complementary/alternative treatments, and medication programs.

Moreover, the classification of mental health services in the Colombian healthcare system is divided into three levels. First (I) level (low) for basic ambulatory care principally in small municipalities, second (II) (medium) for specialised HCPs with medium-range medical technology and third (III) (high) with the highest level of technology and subspecialised personnel (46). Likewise, the coverage was considered local (small municipalities), regional (a whole department/state) and national.

Initial data extraction was performed on a first version with two studies to verify the practicability of the extraction form. After some adjustments, the extraction was completed with all the selected studies. The CFIR was adapted based on the nature of these interventions and the reported information. Therefore, all the domains except for one (individual characteristics) were included. Lastly, information was incorporated using a narrative synthesis method, which facilitated the categorisation of synthesised data and their organisation in text categories (47).

## Results

### Study selection

The search yielded a total of 1 530 results. A duplicate detection process was performed in two phases. Firstly, the detection was supported by Mendeley’s software tool to check duplicates. Based on a reported accuracy of over 95%, 250 duplicates were removed. Finally, 135 possible duplicates were manually examined, finding that eight results were not duplicated; therefore, an additional 127 duplicates were removed.

1 153 results were imported on Rayyan for the title and abstract screening, of which 32 studies were selected for a full-text review, applying the full inclusion and exclusion criteria. 15 studies were excluded due to the lack of description of mental health services (48–62), two because of their focus on characterising mental health disorders (63,64), two were only recommendations and were not implemented (65,66), and 1 due to the type of publication (67). In the end, 12 studies were selected for data extraction. The screening and selection process is presented using the PRISMA flow diagram (Fig 1) (68).

**Figure 1.**
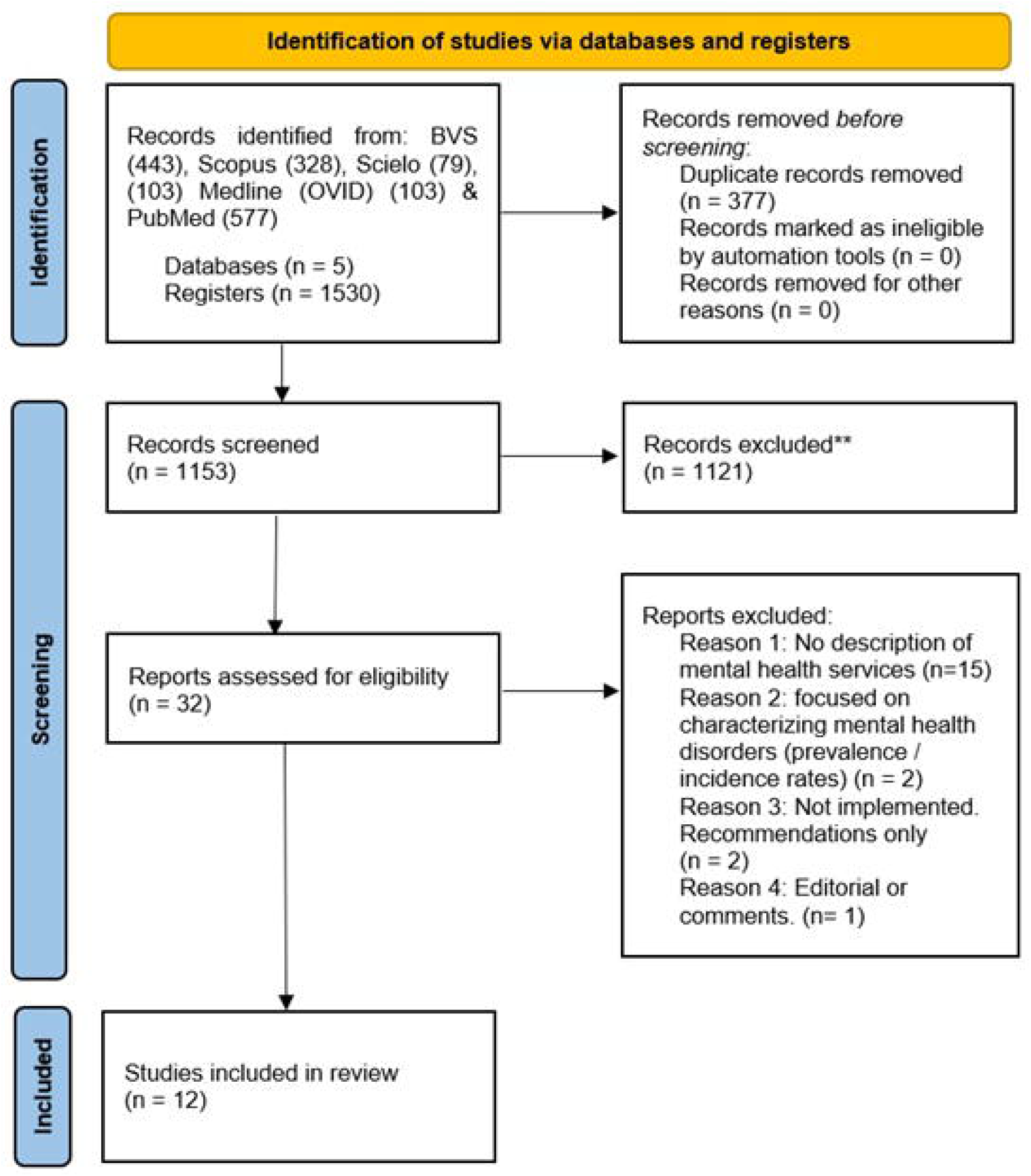
PRISMA – Study selection process.

### General features of included papers

The studies were published in the last six years. Of the 12 selected studies, five were focused on substance use disorders, specifically heroin and alcohol consumption (69–73); three addressed mental health disorders such as anxiety, depression, PTSD, schizophrenia, and bipolar affective disorder (74–76); and the others were centred on suicide (77), behavioural health disorders (78,79), and inclusion and rehabilitation (80).

Geographically, there was high variability since only four studies were solely conducted in the principal cities of Bogota or Cali, while the rest were in multiple smaller cities and municipalities in the country, mainly in rural areas. Notably, three studies were multicentric, combining both urban and rural settings. Similarly, the most reported geographic level attention was local, which means that service is delivered by smaller settings focused on specific districts or neighbourhoods, different to regional and national levels that cover wider populations (7).

Ten studies reported the complexity level of the healthcare institutions, of which seven provided healthcare at the low and middle levels. Likewise, it was significant that all the studies involved multidisciplinary HCPs. Mainly, general practitioners, psychiatrists, psychologists, nurses, and social workers participated. Nevertheless, five studies included infrequent providers such as physical education instructors, nutritionists, occupational therapists, schoolteachers, and even internally displaced women who were trained as part of a program. Four studies were classified as complementary/alternative treatments, four as psychotherapy and four as medication programs along with outpatient or inpatient treatment.

Finally, about the target population, half of the studies were directed to the general population (adults or children), while the rest pursued only men, IDW, university students or users of psychoactive substances.

### Quality of evidence

The studies’ quality was assessed using the Mixed Methods Appraisal Tool (MMAT). This tool was selected because it is feasible to evaluate mixed studies reviews (81).

Overall, the studies had a quality performance of 3.6 over 5.0 on average, with the lowest and highest quality scores of 2.5 and 5.0, respectively. Seven of 12 studies scored above 4.0, and none below 2.5. Notably, lowest scores revealed issues with sampling strategy and coherence between data, methods, and discussion. Also, half of the selection were quantitative descriptive studies, while the other half were qualitative. Further information is provided in **S2 Appendix 2. MMAT scores**.

### Findings

We report on our findings using the broad structure of the CFIR (constructs and domains) complemented with the selected factors of the Socio-Political Economy of Global Mental Health framework.

## 1. Intervention’s characteristics

### Intervention source

Mental health interventions were developed and implemented via partnerships between universities, healthcare providers, and government bodies. Studies also involved international research institutions from North America (Canada and The United States) (17,74,82,83).

### Evidence strength & quality

Studies were supported by primary systematic reviews, national public surveys like the National Mental Health Survey in 2015 (83,84) and previous international studies with notable performance results in similar settings (82,85,86).

### Relative advantage

All the reviewed alternative interventions showed significant advantages in comparison to standard treatment options. One intervention reduced access barriers like geographical isolation (84), treatment continuity, and HCP and patient satisfaction. Another positive point was the approval from settings administrators and will to support implementation, evaluation and continuity (71,75,80). Nevertheless, the reported disadvantages were low articulation with pre-existing service models and no existence of infrastructure and skilled staff (83,87,88).

### Design quality & packing

Half of the selection did not report any toolkit, online or onsite materials for the intervention. The most common resources included prearranged support material from the international sponsor organisation (82,83) and audio-visual materials and software for monitoring and evaluation (84,89).

### Cost

It was remarkable that only two studies referred to financing sources in the interventions being evaluated. The study from Satizabal-Reyes and Ortiz-Quiroga (2020) (80) clarified that no public funding was involved in the program and its continuity merely relies on the urgent and accurate implementation of the National Mental Health Policy. By contrast, in the study by González et al., (73) 43% of the patients paid for their treatment and were treated by private providers, while the remaining patients were covered by the healthcare system and attended public facilities.

## 2. Outer setting

### Patient needs & resources

Primarily, the interventions were developed considering patients’ thoughts and points of view. Also, patients’ families were both formally and informally consulted. This helped managers understand the population’s needs and patients to consider themselves as a person and not be labelled as a disease or condition. This was verified using evaluation components, informal chats and semi-structured interviews (87,91).

### External policies & incentives

It was notable how the interventions were considered and tailored to the Colombian mental health system. All of them mentioned the 2013 National Mental Health Policy and similar regulations and are aimed to support their application.

Moreover, healthcare facilities had to comply with the national regulation to maintain the services enabled or receive high standards certification, thus sustaining public and private funding (17,83,84,87).

## 3. Inner setting

### Structural Characteristics

There was a unanimous position favouring better coordination with government bodies, healthcare providers and research institutions. Additionally, HCPs’ training opportunities and workload imbalance were aspects in need of improvement.

For instance, the lack of skilled HCPs and the excessive workload for professionals were constantly mentioned. For example, nurses were reported to work more than four times the shift time per week compared to psychiatrists (85,92).

### Implementation climate and tension for change

HCPs welcomed technology-based strategies, but there was some resistance and a need to redefine roles when adapting to new healthcare delivery models (e.g., primary healthcare versus traditional clinical treatment) (17,83,84).

In addition, community-based interventions and programs led by universities or research institutions were more likely to have a favourable implementation climate (87,93).

The studies on opioid use disorders indicated a social opposition to any initiative associated with psychoactive substances. The Colombian armed conflict association with narcotraffic caused a social belief of Colombia being a producer country and not a consumer one. Therefore, it is necessary to increase the collaboration between insurers, providers, researchers, policymakers, police bodies and law entities such as The Ministry of Justice. Hence, Colombia will address the emerging consumption phenomenon and its consequences (85,88).

### Available Resources

Not all the studies reported the accessibility of resources for implementation and replication. The access to resources varied considering the type of institution and the established agreements with healthcare administrator companies. The better the cooperation was, the sooner secure the resources were secured, and the more straightforward implementation was (17,82,88,92).

### Access to Knowledge & Information

The reported programs required diverse types of training. The training varied depending on whether the setting was urban or rural. While urban centres had more training and development chances, rural ones struggled with adequate training resources due to deficient internet infrastructure. Alternatively, the strategy from Ceballos et al., (2016) trained non-HCPs people who were also affected by the problem to be addressed. This intervention was closely supervised and approved by remote and onsite experienced trainers.

The interventions sponsored by international institutions demonstrated more precise and constant training and followed up (17,82,83,86).

## 4. Process

### Planning

Generally, the interventions had approved protocols, guidelines and plans that described phases, strategies, and activities. This eased implementation, prevented avoidable barriers and emphasised the importance of the programs.

### Engaging

Only a few studies stated that outside actors backed them. Supporters included police forces, students, and official health officers (77,85,91,93).

### Executing

Despite the existence of plans and protocols, some implementation difficulties were mentioned. The difference in nature and financing source between public and private institutions was a problem when executing the interventions. Public institutions were more likely to be involved than private ones because the public felt more responsible for the health status of their community (17). Nevertheless, public centres presented more access barriers and less retention of patients.

Consulted HCPs reported that time was a frequent problem during execution because the time consumed during piloting, implementation and evaluation was not considered in their workload. Both situations could be certainly addressed with better management practices (83,88). Nonetheless, half of the selection stated that implementation was conducted as planned with minor anticipated obstacles, such as technical and logistical difficulties.

### Reflecting & Evaluating

Although implementation challenges were reported, most of the selected studies reported positive results that were confirmed after assessment procedures. Two interventions developed their evaluation forms (84,93), whereas others utilised established tools commonly used worldwide (17,82,86,88). Information collected on these instruments was individual, assessed performance, and compared indicators against pre-existing programs before and after completion.

On the other hand, two studies did not report assessments because of the nature of the studies, which mainly focused on show characteristics, advantages and development rather than their evaluations (77,87). Only one study reported that an impact evaluation would be performed in the future considering the long-term design of the intervention, which was focused on preventing violence in children and young people using a positive promotion approach (91). Hernández Holguín et al., (78) followed this approach based on the public health ecological model to implement strategies in different spheres such as family, school, and neighbourhood, where children and parents were supported in topics such as nutrition, routines, and crying.

Methodologically, quantitative studies supported their results through descriptive statistics, characterising patients, and explaining programs’ characteristics and treatment models, among other variables (17,82,84,85,88,92). Alternatively, qualitative studies reported participants’ viewpoints, inviting them to provide their perspectives about the programs’ performance and feedback (83,94,95).

## 5. Socio-political economy in mental health

### Gender and Sexuality

Overall, interventions mentioned words such as “female or male” only for descriptive statistical purposes. There was no mention of gender, LGBT (Lesbian, Gay, Bisexual, and Transgender), hetero or homosexual, or other related terms.

One study highlighted that women and their children affected by conflict were often forced to displace to suburban locations where they faced socioeconomic hardship and stigma. Considering this situation, the intervention was focused on women, because improvement in their mental health could impact the mental and physical health and functioning of the household (74).

Another study reflected on reviewing the gender perspective in terms of parenthood, contemplating differences in caregiving according to child sex. This perspective aimed to shape more suitable strategies for preventing violence and promoting positive development for children and young people considering gender roles (78)

### Racism, Caste & Xenophobia

Associated terms such as race, racial, ethnicity, or xenophobia were not mentioned in the selected literature. Only one study acknowledged culture and nationality as an influence on the intervention’s performance. Shannon et al., (71) stressed how interventions and educational material need to be culturally appropriate and deem social influences that tend to lead to exclusion or practices based on preferences. All this increases responsiveness and involvement, impacting positively mental health outcomes.

### Age & Disability

One study mentioned older adults in their analysis, exploring the main reason suicide in this group was related to structural social and economic conditions within specific historical periods. This study suggested that enhancing the social and institutional response to these conditions might impact positively mental health factors that go beyond the psychologist and psychiatrist’s scope (77).

Three studies mentioned how mental health illness was a factor in disability, and deteriorating quality of life in both the individual and their family and community (70,71,80). Substance abuse was a major issue in increasing mortality and contributing to the rising of years lived with a disability (YDL) and Disability Adjusted Life Years (DALYs) (96)

## Discussion

This review identified the mental health services in Colombia assessed in studies and published in peer-reviewed journals. It provided background information about the history of mental health services and the healthcare system, and it explained how this has shaped modern models of service provision. Finally, it has assessed implementation processes using a modified CFIR with the Socio-Political Economy of Global Mental Health framework by Burgess et al., (43) to make it more sensitive to wider social and political factors known to shape the Colombian context.

### Identified mental health services

The first objective of this review sought to determine the mental health services established in Colombia as reported in peer-reviewed literature. Reported interventions offered a broad overview of how mental health has been considered and changed in the last years.

The compiled information showed that the studies focused on certain common and severe mental health conditions such as substance abuse, anxiety, and schizophrenia. Only one study was focused on inclusion and rehabilitation strategies. The types of interventions were psychotherapy, medication programs and complementary treatments. The target population was diverse, with a high majority focusing on general adults and children and specific groups such as users of psychoactive substances and armed conflict victims. Furthermore, studies were broadly completed in some small municipalities and towns where sometimes even essential healthcare services like general practitioners or laboratory services are unavailable.

### Community mental health

Prior studies have noted the importance of rethinking mental health services and treatments within a more coordinated mental health system. This system ought to act concertedly with reasonable opportunities for all the population, but with different approaches to special populations such as armed-conflict victims, women, and LGBTQ+ groups, among others(74,77,79,80).

A tendency for more community mental health involvement was observed in the results. As other authors and health agencies have suggested, a community-centred mental health approach promotes equity and increases access to community resources and patient satisfaction (97,98). Studies from this review align with worldwide evidence and positive experiences of holistic approaches and social recovery rather than biomedical models and clinical recovery (99,100). These studies also recommend disseminating this approach and strengthening existing community mental health services (69,70,80,101). Ultimately, it is recommended to continuously assess implementation as a whole process and complement it with impact evaluation. As previously emphasised by other authors, impact evaluation is vital for global mental health. Measuring factors such as access, coverage, costs, and quality facilitate influencing policymakers to prioritise mental health interventions and strengthen the relevance of mental health interventions (102)

### Challenging psychiatry hegemony

Seven studies were conducted in healthcare settings of low and medium levels of complexity, possibly not requiring an inpatient regimen and including multiple HCPs.Surprisingly, all the interventions included multidisciplinary teams during implementation, contrasting with the traditional psychiatry hegemony.

Historically, in Colombia, mental health services were primarily focused on clinical understandings of mental health, thus contributing to the medicalisation of human life (60), which promotes the excessive medical control of non-medical affairs and the overlooking of social factors instigating disease.

Not instituting mental health services in psychiatric discourses was a relevant tendency identified in this review. The use of multidisciplinary professionals gives different perspectives on mental health and expected mental health outcomes and prevents the repetition of harmful medical treatment overuse such as incarceration, electroconvulsive therapy, and lobotomy (103). This finding is consistent with previous works about the need to reject hegemonic processes in global mental health, preventing the re-colonisation of mental health (104,105), and replacing the mental health practice with strategies that do not follow pathologisation and pharmaceuticalisation processes. Previous works in Brazil have associated these processes with a public health crisis, such as the uncontrolled use of psychotropics in vulnerable settings where social and economic factors are relevant but not deemed (106).

In addition, and following this trend, services were active in considering the need of users, families, and other stakeholders. These diverse perspectives mainly were welcomed by managers because they offered more quality and order. Organisations and providers were open to hearing patients’ concerns and building together treatment options. This attitude is consistent with multiple recommendations for mental health services to follow the recovery approach, empowering patients to participate in their treatments actively (107), understanding that services might differ for specific groups (108) and considering the SDMH comprehensively(100).

### Implementation of mental health services

Services were characterised by being evidence-based, guided by the 2015 National Mental Health Survey and previous clinical studies. This approach strengthened the potential advantage and success of comparing it with standard services.

Studies rarely reported costs associated with implementation. This could be explained due to the financing nature of the reported services (most of them funded by non-public or governmental sources), so the budget may not be open-source information. Nevertheless, this information may be found in other sources like budget reports that were out of the scope of this review.

Though positive, this determination should be interpreted with caution because it may be motivated by funding attraction or certification and enablement process instead of genuine interest in changing the delivery model. Nevertheless, with proper regulation and supervision, this is a remarkable advancement in mental health services centred on people’s needs.

Another important finding was the level of international cooperation as a support mechanism. For many years, overseas institutions have advocated mental health initiatives in Colombia during all their phases (69,71,74). While some institutions backed services with mentoring and training, others provided grants and research scholarships. Governmental support is relatively limited to allow data access and use of infrastructure instead of economic funding (76,80).

Some internal characteristics within organisations facilitated implementation. HCPs, staff, and users embraced the use of technology for service delivery. This facilitated the implementation of telemedicine services, also called telemental health (109), which prevented geographical barriers to access without jeopardising the quality and relationship between users and HCPs. Different Colombian experiences have shown the advantage of promoting these services to address mental health disorders.

Sánchez Díaz et al., (66) highlighted the practicality of telemental health, in particular phone calls to follow-up patients where mental health professionals are not available, while Torrey et al., (110) underlined that the use of telematic tools for screening increased the number of early diagnoses of depression in primary care services where it might be easier to address.

Although telemental health is a strong tendency with multiple advantages for providers, users, and HCP, it should not be used as a replacement strategy for face-to-face services but as a complementary strategy. If necessary, patient needs ought to be reviewed and addressed accordingly with specialised services. Virtual interactions should be used in individual situations where it might be beneficial in comparison (e.g., group interventions, patients with physical disabilities or geographical difficulties in accessing services) (111).

It is notable that even when telematic services are thought to enhance mental health services coverage, they might have their own access’ barriers when patients lack technological access and expertise (111). They could also be unsuccessfully implemented due to clinicians’ and managers’ lack of trust in efficacy, integration into mental health systems and missed in-person experience (112). Nevertheless, multiple experiences in both HICs and LMICs have underlined how telemental health services might be time and cost-saving and offer flexibility to both patients and clinicians (e.g., no travel hours, fewer missed appointments), which could increase access and coverage, leading to address inequalities in mental health services (113–116).

Resistance to adoption was less when recognised institutions like universities guided the use of technology. Community-based interventions function better when students are involved, particularly students from different disciplines because students facilitate adoption (80). This also accords with other experiences of mental health services implementation in Latin America, where students are highly recognised also by staff and other HCPs (117). Though they are not external actors because they are involved during the entire process within the organisation, it can be said that they champion the process and bridge providers with users positively.

In contrast, implementation was usually disrupted because of the lack of coordination between healthcare actors. Though some authors believe that better management could address this problem, it might require redirecting government bodies’ role in planning, delivery, and regulation.

One example of this was the study by Barrios-Acosta et al., (77) about the institutional response of universities to suicide in students, which was utterly unconnected to government services. Nevertheless, the reported services constantly contemplate national roadmaps and guidelines as crucial references, and they follow them strictly.

In this case, institutions designed their strategies to address suicide, considering national and local regulations but lacking cooperation and communication with external actors. This caused, as in other mental health services, overlapped efforts and reduced efficacy. It is, therefore, usual that services aim to suit the health policy instead of proposing different structural approaches from their planning. This finding was also reported by Aceituno et al., (118) concerning poor coordination and time constraints as factors that impacted the implementation of early psychosis services in Chile.

As in other types of health care, there is still a disparity in workload. There was a considerable difference in shift times between nurses and psychiatrists. Gomez Restrepo et al., (70) associated this difference with not specified costs since hiring psychiatrists for extended periods represents elevated expenses. Unfortunately, this imbalance may create an avoidable hostile labour environment. Likewise, training and execution times were sometimes not considered in the staff workload, creating a reluctance to participate. Although there was no specific information about the relationship between staff salaries and inadequate performance in selected studies, multiple global studies in the last 30 years have highlighted this as a critical issue in implementation (119,120).

Services generally had good planning strategies, such as preliminary agreed plans on which responsibilities and deadlines were clear. Planning facilitated implementation by preventing easily avoidable problems.

Precise design and assembling promote accessibility for users, facilitating adaptation and future implementation (121). However, regarding how the interventions were designed and presented, many did not report toolkits or material used to support implementation. Therefore, it is suggested to highlight the usage of this material since it is also a key step in promoting the dissemination and introducing innovative interventions in new settings (122).

During implementation, the difference between public and private institutions was more evident. It may be related to sources’ availability or nature, but private institutions tended only to have slight difficulties. This trend contradicts previous studies in low-and high-income economies (Nepal and Australia), where settings were also influenced by private organisations but shifted to mixed partnerships and independent public-funded services in the last years (123,124).

Australian experience suggested prioritising the allocation of public funding to public mental health services rather than private ones because this support can result in better-positioned and strengthened mental health systems (123). In contrast, the Nepal case proposed improving public and private partnerships, including a comprehensive model where diverse partners are involved. Both Nepal and Colombian cases were generally in low-resource settings and were consistent with the limited inclusion of other partners different to the usual overseas institutions and public settings.

Unlike Colombia, the Nepal study recommended the participation of non-governmental organisations, healthcare service users, multicultural professionals and academic medical centres (124). These examples are feasible options and are in line with previous global guidelines to develop infrastructure for mental health financing, where elements such as involvement of stakeholders, better information systems and evaluation analysis (125).

Finally, regarding evaluation processes, these were performed using designed evaluation forms and validated instruments. This strategy allowed institutions to identify weaknesses, improvement opportunities and future approaches to secure implementation achievement. Unlike other Latin American settings, where the evaluation was utterly requested to participants and non-participants (70), most of the studies in Colombia did not request feedback from all actors (patients, administrative staff, or providers) but just from patients.

This situation can lead to incomplete quality improvement plans and inadequate reflection of organisational priorities (126).

### Socio-Political interactions in mental health

Overall, selected studies were ambiguous in considering the influence of gender, race, and age, in undermining people’s mental health.

Regarding gender and sexuality, the studies limited the use of related terms for statistical purposes. Only 3 studies reported reflections regarding the influence of gender on mental health (71,74,78). For example, Shannon et al., (71) identified that patients and providers were conscious of the gender differences in depression and treatment seeking. In fact, women were more often willing to know more about symptoms and seek out support for depression, while men were more reluctant in accepting symptoms, diagnosis, and treatment due to the *“machismo”* culture (71).

Previous experiences in Latin America have unveiled the relevance of gender perspectives in mental health models and the particular influence of the “machismo” and the less-known “marianismo” on it. Marianismo is the influence of catholic beliefs in Latin women, mainly related to the importance of the Virgin Mary paradigm, normalising women in their submissive feminine role based on their greater moral and spiritual strength (127)

In lived experiences and peer-reviewed studies (128–130), both gender role expectations have been associated with detrimental emotional health outcomes (depression and anxiety symptoms, or violent and hostile behaviours) that are also linked to socio-demographic factors, mentioned before (e.g., race, age, nationality). In addition, the effects of these expectations are particularly worse in LGBTQ+ people due to homophobia, racism and fetishization (129)

Second, though discrimination and stigma based on race and ethnicity are two key risk factors for mental illness (43), none of the studies referred to this, only one work underlined the importance of adapting material and educational strategies based on factors such as ethnicity and cultural viewpoints. Multiple works have enunciated race and ethnicity as social determinants that preferentially disadvantage certain groups in numerous social matters, such as healthcare, marginalising them and affecting their well-being (131–133).

Finally, although age is an influence that might lead to discrimination or stereotyping selected (43), studies did not consider it a relevant factor for their interventions. Only Barrios-Acosta et al., (77) highlighted how suicide in older adults is frequently associated with structural conditions, but this is not deepened in the study, which is focused on young university students.

In line with Flores et al., (134) and Giebel et al., (135) there is scarce literature on older people and mental health in Colombia and only a few studies have explored this field. For instance, Flores et al., (134) used quantitative data which associated higher levels of depression in older adults with earlier armed-conflict situations such as sexual abuse and displacement. However, as in our selected studies, these work and previous ones have failed in incorporating qualitative experiences of life events on peoples’ mental health, which might broaden and analysis and debate regarding mental health services and policies.

Regarding disabilities, only Taborda-Zapata et al., (75) commented on how the answer of the traditional mental health model has been towards hospitalization, prolonging hospitalizations that have led to direct access to institutionalization, thus prolonging disability.

In general, the selected studies did not grapple with the relevance of the selected constructs in mental health. The inclusion of gender, race or age/disability perspectives in mental health discussions in Colombia is still unsettled. As previous studies have endorsed (136), it is suggested for future mental health research to string together socioeconomic factors that shape mental health and well-being and not limit the discussion towards institutional performance based exclusively on indicators. It is necessary to explore expanded viewpoints in people from LMICs, who face certain social and political circumstances. This comprehensive understanding might lead us to recognize the wider impact of these factors on the country’s goals of achieving healthy relationships and well-being.

### The evident but missing association with armed conflict

Lastly, although Colombia has been one of the most affected countries by long-term armed conflicts for the last decades, the included studies were vague on linking conflict and mental health. 11 studies did not mention this major factor, while 1 merely mentioned it only in the background section (74)

Earlier works have stressed the influence of conflict-related violence on people’s mental health. Chaskel et al., (137) described how armed insurgency and internal displacement lead to increased substance misuse, disability, and suicide. Also, Cuartas Ricaurte et al., (138) analysed the relationship between exposure to the armed-conflict and violence with mental health disorders in Colombia concluding that exposure to violent crimes multiplied substantially by socioeconomic difficulties and the overall risk of mental health disorders. This relationship is in line with previous studies in Colombia that also linked violence with socioeconomic deprivation in terms of unemployment and poverty, and together with lack of assistance from the government lead to mental health disorders (139).

### Strengths and limitations

The results from this review may be used for the public debate about the nature of mental health systems and services in Colombia and how they are accomplishing or failing to people’s mental health. To our knowledge, it is the only compilation of this information, including positive changes and the efforts to include the SDMH in policies, systems, and services in Colombia.

To guarantee the consistency and transparency of this systematic review, we followed the PRISMA guidelines for reporting and applied the MMAT to assess the quality of the included studies. However, one limitation was that data review and extraction had no second reviewer to discuss conflicts and reduce bias.

The main limitation of this study was the paucity of detailed publications depicting the implementation of mental health services. Despite applying a comprehensive search strategy covering all the databases deemed relevant, only twelve papers were found to describe a mental health service or intervention implementation. An added related factor is that these services are possibly explained in grey literature from governmental or non-governmental sources. Therefore, not having included government documents and policies or grey literature reduced the generalisability of assumptions about the availability of these services in the country.

## Conclusion

This review has reflected on mental health services in Colombia and their implementation barriers and facilitators. Even though data revealed that mental health services involved strong multidisciplinary teams and extensively adopted the idea of considering users, communities and stakeholders’ viewpoints, there is a notable gap between mental health planning and the implementation of services. There is also an evident disregard for social and economic factors such as gender, race, age, and the influence of the long-run conflict in Colombia. Selected studies were distant from these concepts and historical events, and their impact on mental health, mostly only mentioning them to segregate reported results or provide background.

Remarkably, Colombia has started a long-term public health plan to comprehensively address mental health through laws, policies, and strategies. The aim of this plan is to promote mental health in protective environments, prioritising integral treatments and enhancing intersectoral order and integration (140). However, a critical reported barrier has been the lack of coordination between insurers (EPS), providers, and policymakers with negative consequences on implementation and access to quality care. To redress these challenges, it is necessary to restructure governments’ responsibility within the mental health system, avoid overregulation, and give people and communities more voice and control over their lives and health, as evidence has recommended (97)

Findings suggested the potential of implementing modern models in mental health services in Colombia, involving strategies based on communities, users’ perspectives, and emancipation from the biomedical model. Nevertheless, further efforts are needed to adapt these elements to achieve culturally responsive mental health services with expanded access and increased quality. Moreover, it is necessary to continue exploring complex processes such as mental health services, studying their implementation factors, and involving cost information to achieve substantial evidence. Finally, as underlined throughout, it is advisable to persist in adopting community-based mental health services, and the recovery approach and contemplate gender, race and age as influences that affect mental health. In doing so, these may build healthier, more organised, and empowered individuals and communities.

## Supporting information

S2 Data. Appendix 2. MMAT scores

S3 Data. PRISMA abstract checklist

S4 Data. PRISMA checklist

S1 Data. Appendix 1. Search strategy

Table 2 - Inclusion and exclusion criteria

Table 1 - Overview of the CFIR

## Data Availability

All data and related metadata underlying the findings reported are included in the submitted article and its supplementary information files.

