## Supplementary material for "Mental health services implementation in Colombia – a systematic review": S2 Data. Appendix 2. MMAT scores

| Article | MMAT score |
| --- | --- |
| Barrios Acosta et al., 2017 | **** |
| Borda et al., 2021 | *** |
| Castro et al., 2020 | **** |
| Ceballos et al., 2016 | **** |
| Gomez Restrepo et al., 2018 | **** |
| González et al., 2019 | **** |
| Hernández Holguín et al., 2017 | **** |
| Martínez Pérez et al., 2020 | **** |
| Mejía-Trujillo, Pérez-Gómez and Reyes-Rodríguez, 2015 | ** |
| Satizabal-Reyes and Ortiz-Quiroga, 2019 | *** |
| Shannon et al., 2021 | ***** |
| Taborda Zapata et al., 2016 | *** |

The total possible score for each article is *****
