## Supplementary material for "Mental health services implementation in Colombia – a systematic review": S1 Data. Appendix 1. Search strategy

| **Category** | **Detailed keywords and search equation** |
| --- | --- |
| **English** | |
| *Mental health / mental disorders* | ((mental health)) AND ((mental disorders) OR (neurocognitive disorders) OR (substance-related disorders) OR (mentally ill persons) OR (wellbeing) OR (disability) OR (stress, psychological)) |
| *Location* | ((Colombia) OR (Bogota) OR (Medellin) OR (Cali) OR (Cartagena) OR (Barranquilla)) |
| *Search equation* | ((mental health)) AND ((mental disorders) OR (neurocognitive disorders) OR (substance-related disorders) OR (mentally ill persons) OR (wellbeing) OR (disability) OR (stress, psychological)) AND ((Colombia) OR (Bogota) OR (Medellin) OR (Cali) OR (Cartagena) OR (Barranquilla)) |
| **Spanish** | |
| *Salud mental / trastornos mentales* | ((salud mental) AND ((Trastornos Mentales) OR (Trastornos Neurocognitivos) OR (Trastornos Relacionados con Sustancias) OR (bienestar) OR (discapacidad) OR (discapacidad psicosocial) OR (Enfermos Mentales OR Estrés Psicológico)) |
| *Lugar* | ((Colombia) OR (Bogota) OR (Medellin) OR (Cali) OR (Cartagena) OR (Barranquilla)) |
| *Ecuación de búsqueda* | ((salud mental) AND ((Trastornos Mentales) OR (Trastornos Neurocognitivos) OR (Trastornos Relacionados con Sustancias) OR (bienestar) OR (discapacidad) OR (discapacidad psicosocial) OR (Enfermos Mentales OR Estrés Psicológico)) AND ((Colombia) OR (Bogota) OR (Medellin) OR (Cali) OR (Cartagena) OR (Barranquilla)) |
