## Supplementary material for "Mental health services implementation in Colombia – a systematic review": Table 2 - Inclusion and exclusion criteria

|  | **Main aspect** | | **Inclusion criteria** |
| --- | --- | --- | --- |
| Type of documents | | Published peer-reviewed papers or manuscripts where mental health services were mentioned and/or described. | |
| Publication period | | January 1, 2011, to July 01, 2021 | |
| Language of publication | | No restriction | |
| Include policy, programs/interventions/strategies | | That applied or implemented any of the following:   - Mental health services such as:   - Psychiatric hospitalization, psychotherapy, outpatient and inpatient mental health treatment, recovery, and complementary/alternative treatments, medication programs. | |
| Country of focus | | Colombia | |
| Main aspects | | **Exclusion criteria** | |
| Type of documents | | - Manuscripts, files, or reports that are not openly available. - Editorial or comments. - Newspaper articles. - Scoping/systematic reviews. | |
| Type of services, actions, or interventions | | - Mental health services that are not currently working or are due to be discontinued. - Mental health services that were part of development programs but were stopped. - Studies or publications that are only focused on characterizing and/or analyse mental health disorders (prevalence/incidence rates). | |
