## Supplementary material for "Mental health services implementation in Colombia – a systematic review": Table 1 - Overview of the CFIR

| **Domain** | **Definition** | **Selected constructs** |
| --- | --- | --- |
| *Intervention characteristics* | The influence of essential and adaptable interacting components in the success of implementation. | Intervention source  Evidence strength & quality  Relative advantage  Design quality & packing  Cost |
| *Outer setting* | It is the external framework where the entire process resides. It includes economic, political, and social contexts. | Patient needs & resources  External policies & incentives |
| *Inner setting* | It includes structural features of politics and culture, through which the implementation process will be shaped. | Structural characteristics  Implementation climate  Tension for Change  Available resources  Access to knowledge & information |
| *Individual characteristics* | Features of people who actively experiences during the implementation based on their mindset. | Not used in this review |
| *Process* | Group of sub-processes progressing simultaneously at multiple levels within the organization. | Planning  Engaging  Executing  Reflecting & Evaluating |
